# Colchicine inhibits neutrophil extracellular trap formation in acute coronary syndrome patients after percutaneous coronary intervention

**DOI:** 10.1101/2020.04.20.20034025

**Authors:** Kaivan Vaidya, Bradley Tucker, Rahul Kurup, Chinmay Khandkar, Elvis Pandzic, Jennifer Barraclough, Joshua Machet, Ashish Misra, Mary Kavurma, Gonzalo Martinez, Kerry-Anne Rye, Blake J Cochran, Sanjay Patel

**Author notes:** **Addresses for correspondence:** A/Prof Sanjay Patel, Department of Cardiology, Royal Prince Alfred Hospital, Sydney, NSW 2050, Australia, Dr Blake J Cochran, Atherosclerosis and Cardiometabolic Research Group, Department of Physiology, School of Medical Sciences, UNSW Sydney, NSW, 2033, Australia. These authors contributed equally and share first authorship (Kaivan Vaidya, Bradley Tucker). These authors contributed equally and share senior authorship (Blake J Cochran, Sanjay Patel).

## Abstract

**Objective:** Release of neutrophil extracellular traps (NETs) after percutaneous coronary intervention (PCI) in acute coronary syndrome (ACS) is associated with peri-procedural myocardial infarction, as a result of microvascular obstruction via pro-inflammatory and pro-thrombotic pathways. Colchicine is a potent, well-established anti-inflammatory agent with growing evidence to support use in patients with coronary disease. However, its effects on post-PCI NET formation in ACS has not been explored.

**Approach and Results:** 60 patients (40 ACS; 20 stable angina pectoris [SAP]) were prospectively recruited and allocated to colchicine or no treatment. Within 24 h of treatment, serial coronary sinus blood samples were collected during PCI. Isolated neutrophils from 10 ACS patients post-PCI and 4 healthy controls were treated *in vitro* with colchicine (25 nM) and stimulated with either ionomycin (5 μM) or phorbol 12-myristate 13-acetate (PMA, 50 nM). Extracellular DNA was quantified using Sytox Green and fixed cells were stained with Hoechst and anti-alpha tubulin. Baseline characteristics were similar across both treatment and control arms. ACS patients had higher NET release versus SAP patients (p<0.001), which was reduced with colchicine treatment (AUC: 0.58 vs. 4.29; p<0.001). *In vitro*, colchicine suppressed spontaneous (p=0.004), PMA-induced (p=0.03) and ionomycin-induced (p=0.02) NET formation in neutrophils isolated from ACS patients post-PCI, but not healthy controls. Tubulin organisation was impaired in neutrophils from patients with ACS but was restored by colchicine treatment.

**Conclusions:** Colchicine suppresses NET formation in ACS patients post-PCI by restoring cytoskeletal dynamics. These findings warrant further investigation in randomised trials powered for clinical endpoints.

**Graphical Abstract:** 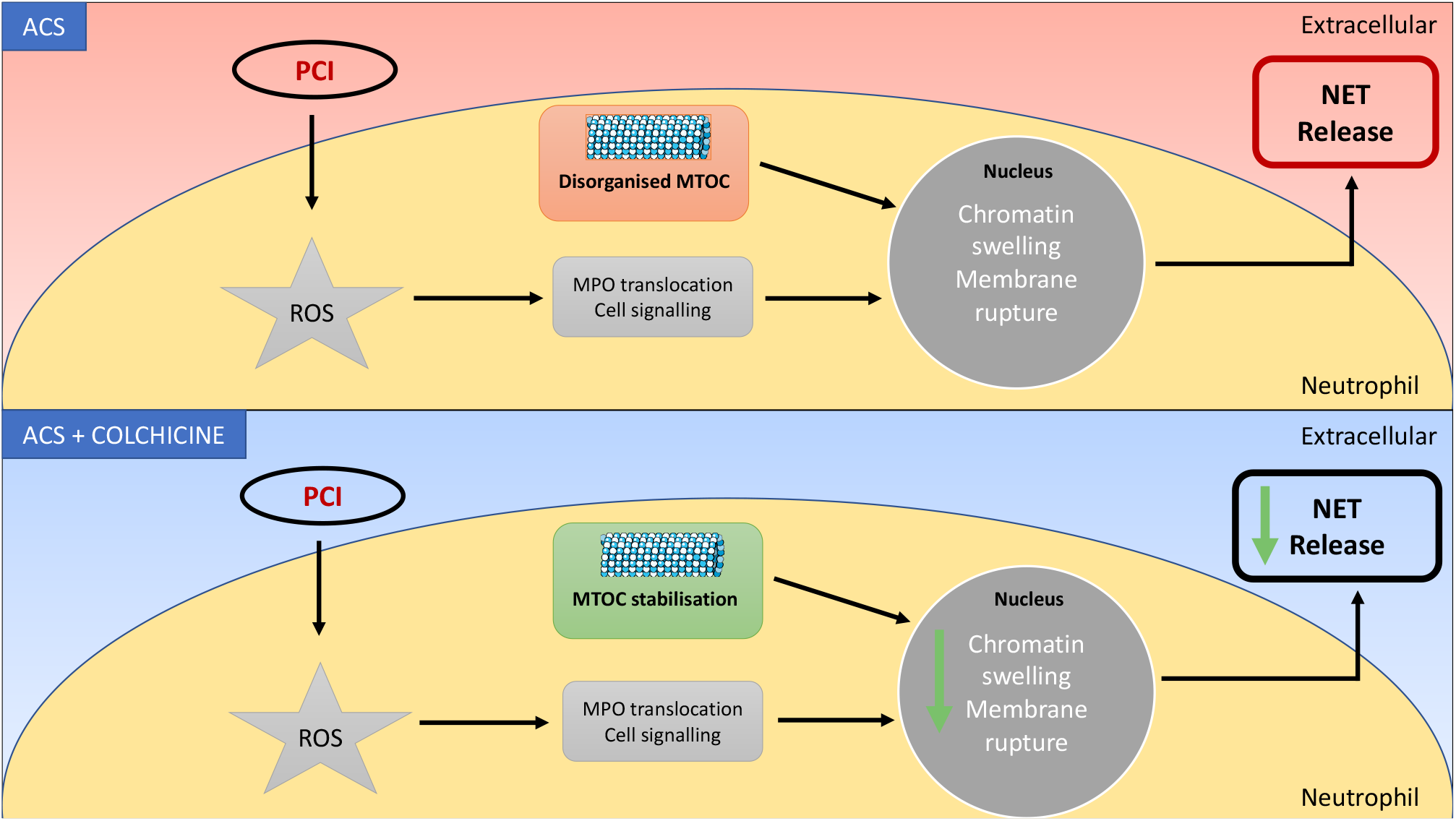

## Introduction

Percutaneous coronary intervention (PCI) in acute coronary syndrome (ACS) patients is associated with a higher incidence of peri-procedural myocardial infarction (MI), due to coronary no-reflow and microvascular plugging by inflamed plaque fragments^1^. This process is believed to be mediated, at least in part, by an over-abundance of activated polymorphonuclear neutrophils in vulnerable coronary plaque that release pro-thrombotic and pro-inflammatory products post-PCI^2^. These products include Neutrophil Extracellular Traps (NETs), a network of extra-cellular DNA, histones, neutrophil elastase (NE), and myeloperoxidase (MPO)^3, 4^. Activated neutrophils secrete NETs via a process known as NETosis, which drives pro-thrombotic and pro-inflammatory cascades by promoting neutrophil-platelet aggregate formation and inflammatory cytokine release from activated endothelial cells, macrophages and T-lymphocytes^3, 5-10^. Classically, NETosis is facilitated by the generation of intracellular reactive oxygen species (ROS) that activate protein kinase pathways and transcription factors, ultimately leading to chromatin decondensation and NET formation^11^. Increased NET release after primary PCI has been shown to correlate positively with infarct size and negatively with ST-segment resolution^3^.

Despite the introduction of high dose statin pre-PCI, peri-procedural MI is still common and affects one in four patients undergoing PCI^12^. It is also associated with a significantly increased risk of in-hospital major adverse cardiac events (MACE) and death^13^. To date, no study investigating the role of a dedicated anti-inflammatory agent in this setting has been published. Colchicine, a well-established anti-inflammatory drug, has emerged as a potential therapeutic tool in patients with coronary disease. Results of the recent LoDoCo^14^ and COLCOT^15^ trials have demonstrated that long-term colchicine treatment significantly reduces the risk of secondary ischemic cardiovascular events. Colchicine treatment has also been shown to promote stabilization of atherosclerotic plaque^16^. Although colchicine inhibits neutrophil chemotaxis^17^, its effect on NETosis, particularly in the post-PCI setting, remains to be evaluated.

Therefore, in this study, we investigated the effects of short-term colchicine treatment in ACS patients and stable angina pectoris (SAP) patients undergoing PCI. We found that it significantly reduced NET release into the coronary sinus of ACS patients, in part via direct cytoskeletal fixation in primed neutrophils, thereby arresting chromatin swelling and NET release.

## Materials and Methods

Please see the Major Resources Table in the Supplemental Materials.

### Patient population

This study was approved by the Sydney Local Health District research ethics and governance office and all participants provided written informed consent. The project was registered with the Australian New Zealand Clinical Trials Registry (ACTRN12619001231134). Sixty participants (40 ACS, 20 SAP^18^) were prospectively recruited from the Department of Cardiology, Royal Prince Alfred Hospital, Sydney, NSW, Australia (Fig. 1A). Patients were allocated in a 1:1 ratio to either no treatment, or treatment with peri-procedural colchicine using a previously described regimen^19^: 1 mg followed by 0.5 mg after 1 h, 6 to 24 h prior to coronary angiography. Patients were allocated to treatment or control groups in successive alternating fashion, with odd numbered patients assigned to receive colchicine from initiation to completion of the study received colchicine. For the *ex vivo* aspect of this study, a further 11 colchicine-naïve ACS patients and 4 healthy controls were recruited (Fig. 1B). Venous blood samples were collected from these ACS patients within 24 h of PCI.

**Figure 1:**
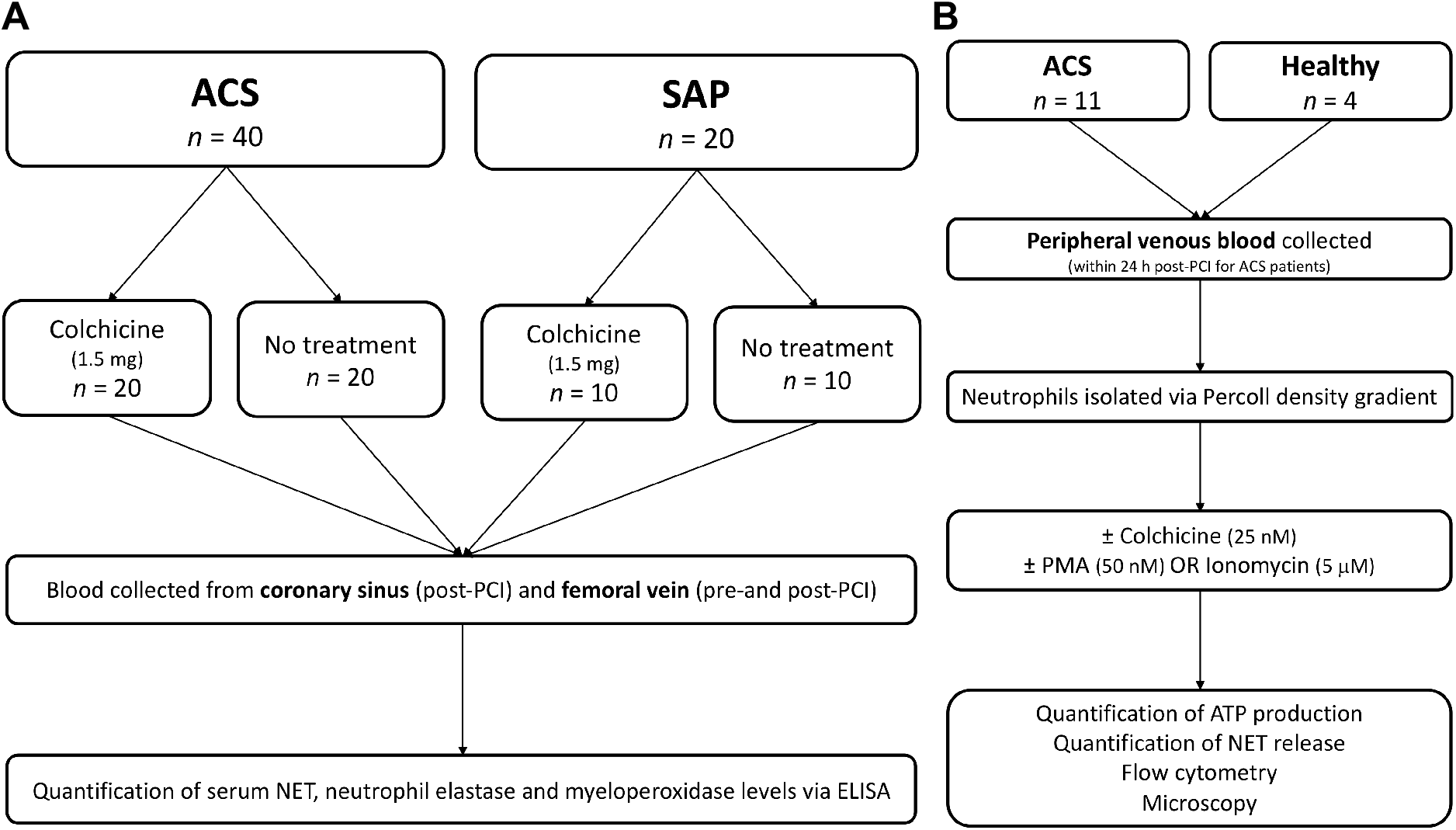
Participant flow. *<u>In vivo</u>* <u>study:</u> ACS and SAP patients were randomised to either colchicine (total dose 1.5 mg) or no treatment. Blood samples were collected from the coronary sinus and femoral vein of patients undergoing PCI for quantification of NET, NE and MPO levels. *<u>Ex vivo</u>* <u>study:</u> Peripheral venous blood was collected from healthy volunteers and patients with ACS (within 24 h post-PCI). Neutrophils were isolated using a Percoll density gradient and incubated with colchicine (25 nM) or vehicle control, followed by stimulation with PMA (50 nM) or ionomycin (5 μM) and assessment of NET release, cytoskeletal organisation, chromatin swelling, ROS production and cell viability. ACS – acute coronary syndrome, MPO – myeloperoxidase, NE – neutrophil elastase, NET – neutrophil extracellular trap, PCI – percutaneous coronary intervention, ROS – reactive oxygen species.

All included patients were over 18 years of age with a clinical indication for a coronary angiogram and PCI. Patients with a >50% stenosis in the left main coronary artery, cardiogenic shock or haemodynamic instability, pregnant or lactating women, known colchicine hypersensitivity, or patients already taking colchicine, were excluded from this study.

### Blood sampling

During PCI, coronary sinus (CS) blood samples were collected at 45 s intervals, as previously described^19^ (Supplementary Table 1). Peripheral venous blood was also collected, pre-and post-procedure from the common femoral vein. CS sampling has been shown in multiple prospective studies to offer an enhanced method of assessing the local cardiac milieu and its associated pathophysiology^20^. Our technique to safely cannulate the CS ostium has previously been described^21^. All patients received unfractionated heparin (70 U/kg). As soon as the activated clotting time was confirmed to be over 250 s, coronary angioplasty and CS sampling was performed. In all cases, stents were deployed at nominal pressure for 30 s, followed by non-compliant balloon pressure dilatation (size and pressure at operator’s discretion).

**Table 1:** Baseline demographic and clinical characteristics.

| Baseline Characteristics | ACS patients |  |  | SAP patients |  |  |
| --- | --- | --- | --- | --- | --- | --- |
|  | No treatment<br>(n = 20) | Colchicine (n<br>= 20) | p value | No treatment<br>(n = 10) | Colchicine (n<br>= 10) | p value |
| Age (years) (n=59) | 66.0 (± 13.4) | 61.1 (±13.3) | 0.27 | 62.4 (± 8.8) | 63.3 (±8.8) | 0.82 |
| Sex (Male) (n=60) | 15 (75%) | 16 (80%) | 1.00* | 8 (80%) | 10 (100%) | 0.47* |
| Presentation (n=60) |  |  | 0.25 |  |  | 1.00 |
| SAP | 0 | 0 |  | 10 (100%) | 10 (100%) |  |
| UAP | 6 (30%) | 11 (55%) |  | 0 | 0 |  |
| NSTEMI | 12 (60%) | 7 (35%) |  | 0 | 0 |  |
| STEMI | 2 (10%) | 2 (10%) |  | 0 | 0 |  |
| Time from presentation to PCI (n=60) |  |  | 0.87 | N/A | N/A | N/A |
| < 24 hours | 2 (10%) | 3 (15%) |  |  |  |  |
| 24-48 hours | 11 (55%) | 11 (55%) |  |  |  |  |
| >48 hours | 7 (35%) | 6 (30%) |  |  |  |  |
| Body mass index (kg/m<sup>2</sup>) (n=57) | 27.7 (±4.1) | 30.1 (±4.6) | 0.11 | 27.8 (±4.9) | 28.9 (±4.7) | 0.60 |
| Blood pressure (mmHg) (n=57) |  |  |  |  |  |  |
| Systolic | 131.9 (±16.5) | 126.3 (±41.1) | 0.85 | 153.8 (±37.2) | 146.5(±29.4) | 0.68 |
| Diastolic | 75.7 (±11.2) | 66.0 (±23.4) | 0.17 | 84.2 (±24.6) | 75.3 (±16.1) | 0.53 |
| Diabetes mellitus (n=60) | 8 (40%) | 11 (55%) | 0.34 | 3 (30%) | 5 (50%) | 0.65* |
| Hypertension (n=60) | 15 (75%) | 12 (60%) | 0.31 | 9 (90%) | 8 (80%) | 1.00* |
| Dyslipidaemia (n=60) | 15 (75%) | 16 (80%) | 1.00* | 9 (90%) | 9 (90%) | 1.00* |
| Positive family history (n=59) | 6 (30%) | 7 (35%) | 0.74 | 5 (50%) | 3 (33.3%) | 0.65* |
| Smoking history (n=60) | 14 (70%) | 12 (60%) | 0.51 | 8 (80%) | 7 (70%) | 1.00* |
| Current smoker (n=60) | 6 (30%) | 7 (35%) | 0.74 | 2 (20%) | 2 (20%) | 1.00* |
| Previous MI (n=60) | 3 (15%) | 6 (30%) | 0.45* | 3 (30%) | 5 (50%) | 0.65* |
| Previous PCI (n=60) | 4 (20%) | 6 (30%) | 0.47 | 4 (40%) | 4 (40%) | 1.00* |
| Previous CABG (n=60) | 1 (5%) | 2 (10%) | 1.00* | 1 (10%) | 1 (10%) | 1.00* |
| Previous CVA (n=60) | 1 (5%) | 1 (5%) | 1.00* | 0 | 1 (10%) | 1.00* |
| Renal impairment<br>(eGFR < 45) (n=56) | 3 (15.8%) | 1 (5.3%) | 0.60* | 0 | 0 | 1.00* |
| Medications (n=56) |  |  |  |  |  |  |
| Aspirin | 18 (100%) | 20 (100%) | 1.00* | 9 (100%) | 9 (100%) | 1.00* |
| Clopidogrel /<br>Prasugrel / Ticagrelor | 17 (94.4%) | 19 (95%) | 1.00* | 9 (100%) | 8 (88.9%) | 1.00* |
| Statin | 15 (83.3%) | 17 (85%) | 1.00* | 7 (77.8%) | 9 (100%) | 0.47* |
| Beta blocker | 14 (77.8%) | 11 (55%) | 0.14 | 6 (66.7%) | 6 (66.7%) | 1.00* |
| ACEI/ARB | 8 (44.4%) | 12 (60%) | 0.34 | 6 (66.7%) | 7 (77.8%) | 1.00* |
| Biochemistry |  |  |  |  |  |  |
| Hs-CRP (mg/L)<br>(n=44) | 21.72 (±29.19) | 15.22 (±24.10) | 0.62 | 3.14 (±1.88) | 1.84 (±1.29) | 0.16 |
| Total cholesterol<br>(mmol/L) (n=47) | 4.53 (±1.83) | 4.82 (±1.43) | 0.38 | 4.03 (±0.70) | 3.96 (±1.23) | 0.71 |
| Absolute neutrophil<br>count (x 10<sup>3</sup>/mL) (n=59) | 4.47 (±1.66) | 5.64 (±2.93) | 0.13 | 3.88 (±1.36) | 4.24 (±0.91) | 0.50 |
| Baseline hs-TnT (ng/L)<br>(n=54) | 554.50<br>(±1011.75) | 882.0<br>(±1639.27) | 0.92 | 12.88 (±11.63) | 15.78(±15.10) | 0.42 |
| **Angiographic characteristics** |  |  |  |  |  |  |
| Diseased vessels no.<br>(n=57) |  |  | 0.03 |  |  | 0.97 |
| One | 7 (35%) | 5 (25%) |  | 4 (50%) | 5 (55.6%) |  |
| Two | 13 (65%) | 9 (45%) |  | 3 (37.5%) | 3 (33.3%) |  |
| Three | 0 | 6 (30%) |  | 1 (12.5%) | 1 (11.1%) |  |
| Coronary artery(s)<br>treated (n=58) |  |  | 0.42 |  |  | 0.65 |
| LAD | 9 (45%) | 7 (35%) |  | 5 (55.6%) | 4 (44.4%) |  |
| LCX | 4 (20%) | 4 (20%) |  | 1 (11.1%) | 2 (22.2%) |  |
| RCA | 7 (35%) | 4 (20%) |  | 3 (33.3%) | 2 (22.2%) |  |
| SVG | 0 | 1 (5%) |  | 0 | 1 (11.1%) |  |
| LAD & RCA | 0 | 1 (5%) |  | 0 | 0 |  |
| LCX & RCA | 0 | 1 (5%) |  | 0 | 0 |  |
| LAD & LCX | 0 | 2 (10%) |  | 0 | 0 |  |
| Stent characteristics |  |  |  |  |  |  |
| Length (mm) (n=45) | 18.1 (±7.4) | 18.1 (±6.8) | 0.83 | 20.1 (±10.1) | 20.1 (±8.2) | 0.90 |
| Size (mm) (n=46) | 2.9 (±0.5) | 2.9 (±0.4) | 0.60 | 2.9 (±0.3) | 3.3 (±0.5) | 0.21 |
| Stent type (n=48) |  |  | 0.06 |  |  | 0.37 |
| DES | 15 (88.2%) | 12 (66.7%) |  | 3 (60%) | 7 (87.5%) |  |
| BMS | 2 (11.8%) | 1 (5.6%) |  | 1 (20%) | 1 (12.5%) |  |
| Bioabsorbable scaffold | 0 | 5 (27.8%) |  | 1 (20%) | 0 |  |
| Lesion type (n=60) |  |  | 0.53 |  |  | 0.87 |
| A | 8 (40%) | 10 (50%) |  | 2 (20%) | 3 (30%) |  |
| B | 12 (60%) | 10 (50%) |  | 6 (60%) | 5 (50%) |  |
| C | 0 (0%) | 0 (0%) |  | 2 (20%) | 2 (20%) |  |
| Corrected TFC (n=60) | 27.5 (±4.7) | 26.9 (±5.7) | 0.62 | 24.0 (±1.7) | 23.8 (±1.7) | 0.97 |

ACEI/ARB – angiotensin converting enzyme inhibitor/angiotensin receptor blocker, BMS – bare metal stent, CABG – coronary artery bypass grafting, CVA – cerebrovascular accident, DES – drug-eluting stent, Hs-CRP – high-sensitivity C-reactive protein, hs-TnT – high-sensitivity troponin T, LAD – left anterior descending, LCX – left circumflex, MI – myocardial infarction, N/A – not applicable, NSTEMI – non-ST elevation myocardial infarction, PCI – percutaneous coronary intervention, RCA – right coronary artery, SAP – stable angina pectoris, STEMI – ST elevation myocardial infarction, SVG – saphenous vein graft, TFC – TIMI Frame Count, UAP – unstable angina pectoris.
\* Fisher's exact test used.

### Blood processing

Plasma was separated from whole blood by centrifugation at 1500 × *g* for 10 min and stored at −80°C. MPO (Hycult Biotech) and NE (Abcam) concentrations were qualified by ELISA. Levels of NETs were assessed using an MPO-DNA ELISA as described previously^22^.

### Neutrophil isolation

Peripheral venous blood was collected from ACS patients up to 24 h after PCI (n=11) and healthy volunteers (n=4). Blood was collected in EDTA vacutainers and neutrophil isolation commenced within 1 h of sampling. Neutrophils were isolated using a Percoll-density gradient, as previously described^23^. This protocol resulted in a granulocyte layer with > 95% neutrophils, as confirmed by surface expression of CD66b via flow cytometry and observation using light microscopy and Romanowsky stain.

### NETosis assay

A plate reader based assay was used to measure NETosis^11^. Isolated neutrophils were resuspended in RPMI medium with 2% (v/v) fetal bovine serum at 5× 10^5^ cells/mL. Sytox Green was added to the cell suspension (5 μM final concentration) and 200 μL of the resulting solution was seeded in 96-well black clear-bottom plates (Corning). Subsequently, colchicine (25 nM final concentration; Sigma-Aldrich) or vehicle control was added to the wells followed by incubation for 1.5 h at 37°C. NETosis was then induced with 50 nM PMA (Sigma-Aldrich) or 5 μM ionomycin (Sigma-Aldrich). Control cells were not stimulated. Fluorescence was measured at 2 min intervals for 4 h using a fluorescence plate reader (504 nm excitation, 523 nm emission; SpectraMax, Molecular Devices). Cells were subsequently lysed using 1% (v/v) Triton-X-100 and fluorescence measured again to determine total DNA content. All values were normalised to total DNA at each time point.

### Cell viability assay

Cell viability was assessed via quantification of ATP levels following stimulation, as previously described^24^. Isolated neutrophils were seeded in white 96-well plates (Corning) at a density of 2× 10^4^ cells/well in RPMI medium with 2% (v/v) fetal bovine serum. NETosis was induced with either 50 nM PMA or 5 μM ionomycin for 0 min, 15 min, 30 min, 60 min and 120 min. CellTiter-Glo Reagent (Promega) was added in equal volume to the cell culture medium present in each well, the plate shaken for 2 min to induce cell lysis and then incubated at room temperature for 10 min to allow stabilization of the luminescent signal. Luminescence was measured (CLARIOstar, BMG Labtech) and values normalized to unstimulated cells.

### Reactive oxygen species

Intracellular reactive oxygen species (ROS) generation was measured using dihydrorhodamine 123 (Invitrogen). Neutrophils (1× 10^6^) were isolated from ACS patients and incubated at 37 °C for 20 min with 5 μM dihydrorhodamine 123 in the presence of absence of PMA or ionomycin as previously described. Fluorescence was quantified by flow cytometry (BD LSRFortessa SORP X-20, Biological Resources Imaging Laboratory, UNSW Sydney).

### Microscopy

Neutrophils were isolated from ACS patients post-PCI and seeded on Nunc Lab-Tek II 8-well chamber slides (ThermoFisher) at a density of 2× 10^4^ cells/well. Cells were incubated for 1.5 h at 37 °C with either colchicine (25 nM final concentration) or vehicle control, stimulated with either PMA or ionomycin then fixed with 2% (w/v) paraformaldehyde for 10 min at room temperature. Cells were permeabilized with phosphate buffered saline (PBS) containing 1% (w/v) bovine serum albumin and 0.3% (v/v) Triton-X100 at 4 °C overnight then stained with Alexa647 conjugated anti-α-tubulin (1:100, Abcam) at 4 °C overnight. Chromatin was stained with Hoechst (1.62 μM final concentration) for 5 min at room temperature. Slides were then mounted on cover slips using ProLong Gold (ThermoFisher) and imaged on PMT (α-tubulin) or Airy detector (Hoechst) using a 63× /14 Plan-Apochromat lens with 405 nm and 647 nm laser lines on Zeiss LSM 880 confocal microscope (Biomedical Imaging Facility, UNSW Sydney).

### Image analysis

Images were loaded in custom written Matlab (Mathworks) scripts. A semi-automatic approach was used to identify individual cells and determine the chromatin area per cell. To characterise the tubulin radial intensity profile from the centre of microtubule organization centre (MTOC), individual cells and the corresponding MTOC were manually selected. For each cell, a circular average of intensity was performed at each radial distance from the centre of the defined MTOC. For each radial intensity profile, the maximum intensity at the MTOC was extracted. Multiple field of views were analyzed for each condition and the average and standard error of mean intensity was calculated at each radius.

### Statistical analysis

Continuous variables were reported as mean (± SD) and categorical variables as number (percentage). The release of NETs, NE, and MPO over 5 sampling time points at 45 s intervals during PCI was quantified by area under the curve (AUC). Results of the Sytox Green NETosis assay were quantified by plotting fluorescence intensity against time and calculating the AUC for each sample. Normality of distribution was tested using the Shapiro-Wilk’s test (p > 0.05) as well as skewness and kurtosis. Differences in continuous variables were tested by unpaired or paired Student’s t test or Mann-Whitney U test, as appropriate. Proportional differences in categorical variables were tested using the Chi-square (χ^2^) test or the Fisher’s exact test for association. A two-tailed p-value of < 0.05 was considered statistically significant. Statistical analysis was performed using SPSS software version 22.0 (IBM, USA).

## Results

### Baseline characteristics

From March 2015 to April 2017, 60 patients were recruited into the study, of which 40 presented with an ACS and 20 had SAP. Baseline characteristics were similar between both the treatment group and controls, with no statistically significant difference for any of the recorded demographic, clinical, or angiographic variables (Table 1) except number of diseased vessels in the ACS patients (p=0.03). The mean age was 63.3 (±11.9) years and 49 (81.7%) patients were male. Of the ACS presentations, there was a higher prevalence of unstable angina pectoris (UAP; 55% vs. 30%) in the treated group and a higher prevalence of non-ST elevation myocardial infarction (NSTEMI; 60% vs. 35%) in the control group. Nearly half of the patients were diabetic (45%), and most had hypertension (73.3%), dyslipidaemia (81.7%) and were on a statin (80%). In the treatment arm, 2 patients underwent vein graft PCI and 4 patients underwent multi-vessel PCI, compared to no patients in the control arm, however CS sampling was only performed during the first (culprit lesion) PCI. Mean corrected TIMI frame count (cTFC) -a standardised, quantitative, objective index of assessing coronary flow^25^ – was normal (≤27) for all patients except the untreated ACS cohort (mean cTFC 27.5 ±4.7). Mean cTFC was higher in ACS patients versus SAP patients (27.2 ±5.2 vs. 23.9 ±1.7), indicating enhanced microvascular dysfunction. There was no significant difference in mean cTFC between untreated and treated cohorts in either group. Finally, the mean Gensini score^26^, a measure of extent of coronary atherosclerosis on angiography, was slightly higher in ACS patients (50.7 ±12.9) versus SAP patients (47.6 ±12.7), with no difference between untreated and treated cohorts for either group.

For the *ex vivo* component of this study, a further 11 ACS patients were recruited from July to September 2019 (Supplementary Table 2). This cohort was not subject to oral colchicine treatment prior to PCI. The mean age of these participants was 71.5 (±14.2) and 7 (63.6%) participants were male. Of the ACS presentations, there was 1 (9.1%) unstable angina pectoris, 7 (63.6%) NSTEMI and 3 (27.3%) STEMI. The majority of these patients were hypertensive 8 (72.7%), with an equal proportion being dyslipidaemic 8 (72.7%). Only 1 (9.1%) patient had diabetes mellitus.

### Events

There were no adverse events related to administration of colchicine, sampling from the coronary sinus, or the PCI procedure itself in any of patients in this study. None of the 20 SAP patients experienced a peri-procedural MI. Of the ACS patients there were 7 (35%) peri-procedural MIs in the untreated cohort and 3 (15%) in the treated cohort.

### Patients with ACS have elevated markers of neutrophil activation in coronary sinus levels pre-PCI

Mean baseline (pre-PCI) levels of NETs (0.205±0.139 v 0.060±0.047 %^22^; p<0.001; Fig. 2A), NE (204.40±54.49 v 132.70±29.04 ng/mL; p<0.001; Fig. 2B), and MPO (164.67±37.08 v 127.25±35.70; p=0.004; Fig. 2C) were significantly higher in ACS patients versus stable controls. There was no significant difference in the baseline values between patients that did or did not receive colchicine (Supplementary Table 3).

**Figure 2:**
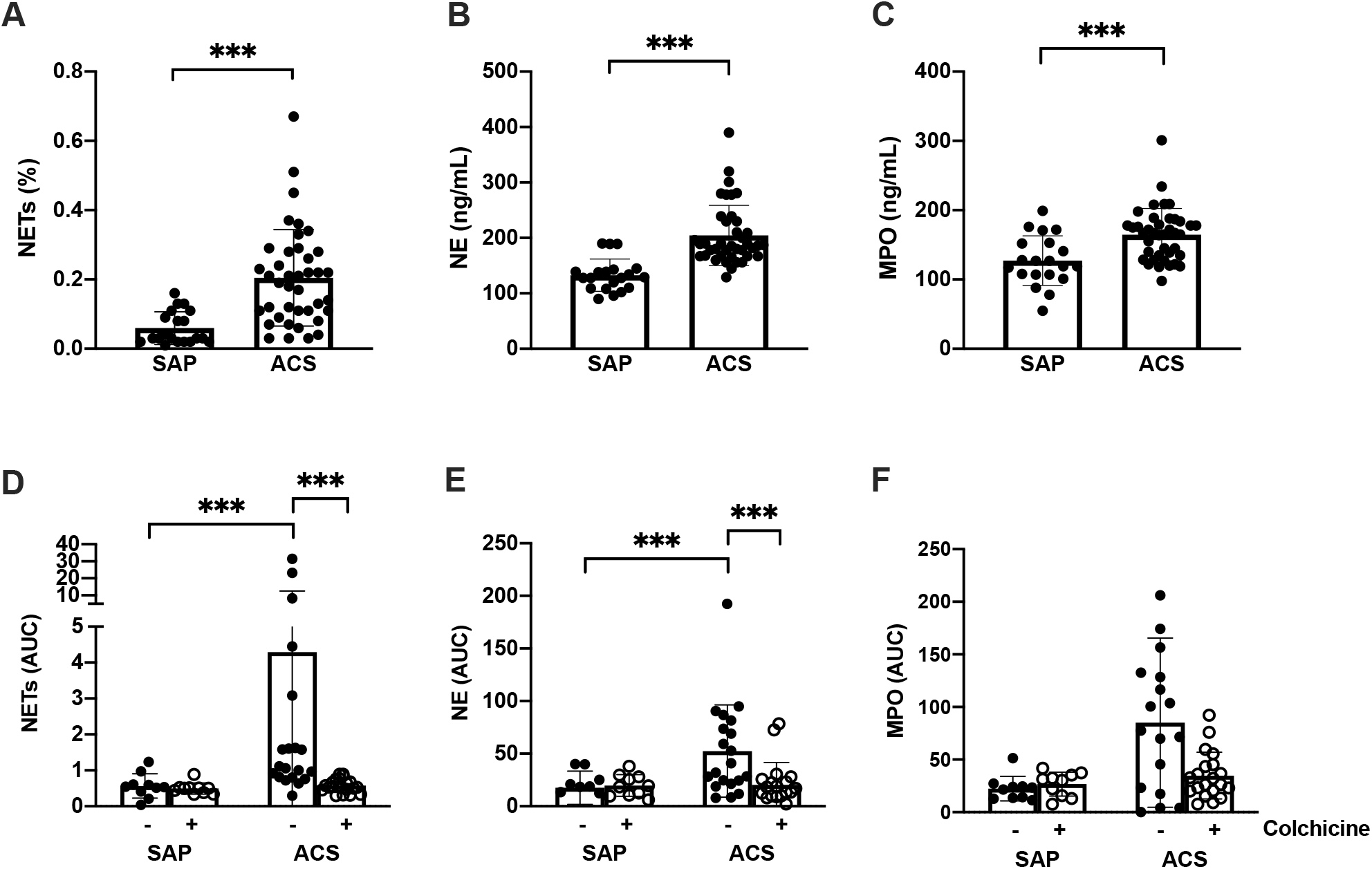
Patients with acute coronary syndrome have enhanced neutrophil activity that is be acutely suppressed by colchicine. Blood was collected from the coronary sinus prior to PCI and levels of (**A**) neutrophil extracellular traps (NETs), (**B**) neutrophil elastase (NE) and (**C**) myeloperoxidase (MPO) quantified. Patients with stable angina pectoris (SAP) or acute coronary syndrome (ACS) were or were not treated with colchicine. Blood was collected from the coronary sinus during the procedure. (**D**) Neutrophil extracellular traps (NETs), (**E**) neutrophil elastase (NE) and (**F**) myeloperoxidase (MPO) levels were measured over time and area under the curve (AUC) calculated. Error bars represent standard deviation of the mean. ***p < 0.005; **p <0.01; *p <0.05.

### Colchicine inhibits neutrophil activation during PCI in patients with ACS

In the untreated patients, both NET (AUC: 4.29±8.17 vs 0.57±0.34; p<0.001; Fig. 2D) and NE (AUC: 52.41±43.86 vs 17.61±15.91; p=0.01; Fig. 2E) levels were significantly elevated in the CS during PCI of culprit lesions in patients with ACS compared to SAP. Whilst MPO levels were also increased, this did not reach statistical significance (AUC: 85.06±80.46 vs 22.42±11.68; p=0.10; Fig. 2F).

Treatment with colchicine resulted in a significant reduction of both NET and NE release in the CS of patients with ACS. NET release was over seven times lower (AUC: 0.58±0.19 vs. 4.29±8.17; p<0.001; Fig. 2D) and NE release was over 2.5 times lower (AUC: 20.27±21.21 vs. 52.41±43.86; p=0.002; Fig. 2E) in colchicine-treated ACS patients compared to controls during PCI. MPO release was also over two-fold lower in the treated patients compared to controls (AUC: 34.75±22.13 vs. 85.06±80.46; Fig. 2F), however this difference did not reach statistical significance (p=0.09). In contrast, colchicine treatment had no impact on the level of NETs, NE or MPO in patients with SAP.

### Colchicine does not impact peripheral levels of activated neutrophil mediators

To evaluate whether release of neutrophil-derived products was a systemic process or localized to the coronary circulation (as measured by CS sampling), and whether colchicine suppressed these enzymes and protein networks, we also compared changes in peripheral venous NETs, NE, and MPO levels (post-PCI minus pre-PCI). There was no difference in peripheral venous levels of all mediators either pre-or post-PCI in both the colchicine treated and non-treated groups. This was true for both patients with SAP and ACS (Supplementary Fig. 1).

### ACS neutrophils are primed for NETosis

To further understand the mechanisms of NET production post-PCI, NET release was measured from neutrophils isolated from both ACS patients post-PCI and healthy participants. In the absence of stimulation, neutrophils isolated from ACS patients displayed increased spontaneous NET formation (AUC: 1.05± 0.60 vs 0.23±0.17, respectively; p=0.026; Fig. 3A). Similarly, in the presence of PMA, neutrophils isolated from ACS patients demonstrated augmented NET formation (AUC: 2.05±0.56 vs 0.88±0.50; p=0.006; Fig. 3B). A similar trend was observed with ionomycin stimulation, although the difference did not reach statistical significance (AUC: 2.41±1.02 vs 1.58±0.69; p=0.18; Fig. 3C).

**Figure 3:**
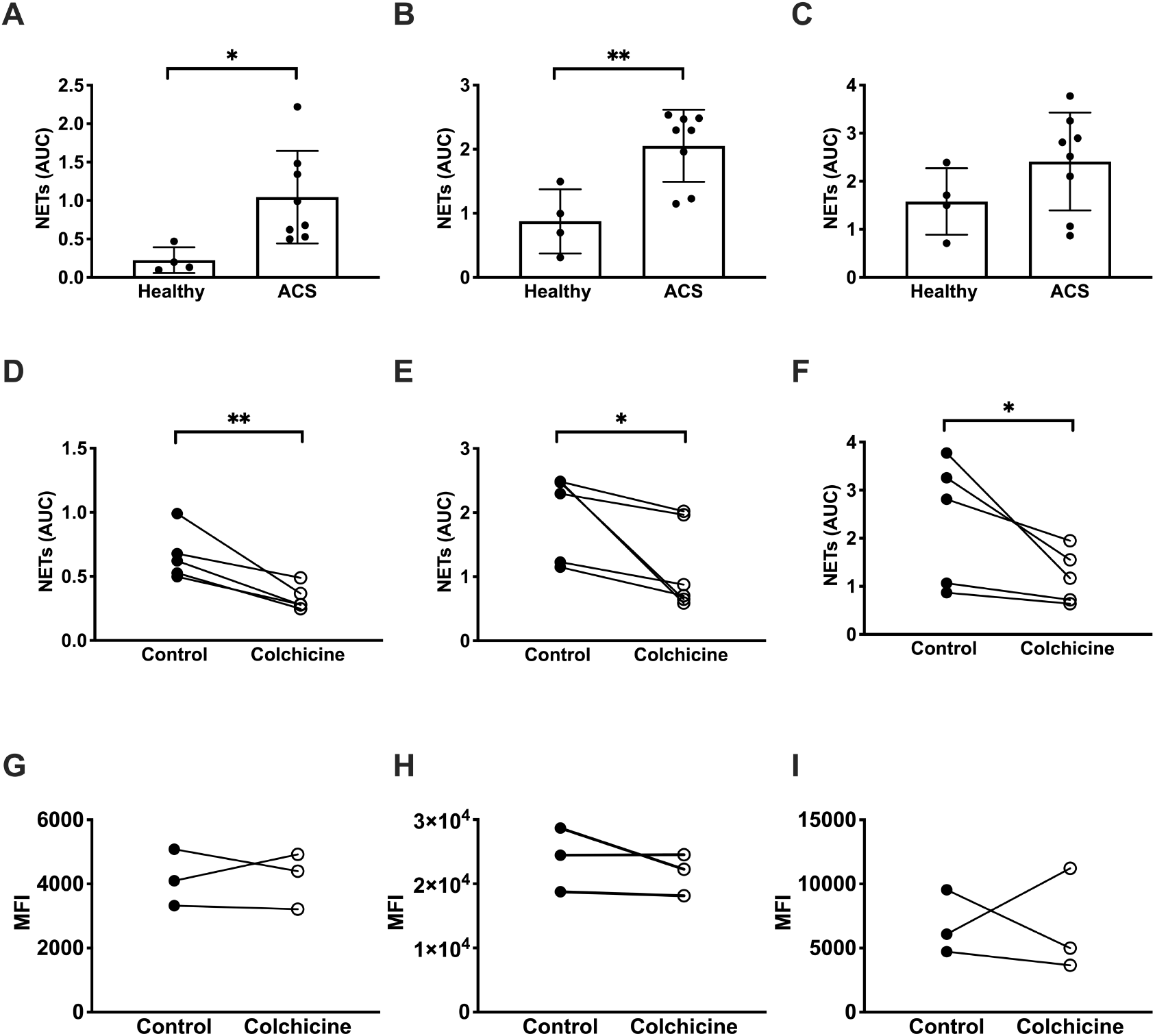
Colchicine directly decreases markers of neutrophil activation in the coronary sinus of patients with acute coronary syndrome during PCI. Neutrophils from ACS patients and healthy participants were isolated and NET release measured using a fluorescent plate-based method in (**A**) unstimulated cells or in cells stimulated with (**B**) PMA or (**C**) ionomycin. Neutrophils from patients with ACS were isolated and treated with 25 nM colchicine for 1.5 h prior to measurement of NET release in (**D**) unstimulated cells or in cells stimulated with (**E**) PMA or (**F**) ionomycin. Neutrophils from patients with ACS were isolated and treated with 25 nM colchicine for 1.5 h prior to quantification of reactive oxygen species (ROS) production using dihydrorhodamine 123 in (**G**) unstimulated, (**H**) PMA-and (**I**) ionomycin-stimulated cells. Error bars represent standard deviation from the mean. **p < 0.01; *p < 0.05.

### Colchicine inhibits NETosis in ACS primed neutrophils

To examine the direct effect of colchicine on NETosis, neutrophils isolated from patients with ACS post-PCI were subjected to colchicine treatment *in vitro* at equivalent physiologic doses. In the absence of stimulation, colchicine markedly reduced spontaneous NETosis (AUC: 0.66±0.20 vs 0.33±0.10; p=0.004; Fig. 3D). Similar results were obtained when isolated neutrophils were stimulated with PMA (AUC: 2.02±0.65 vs 1.13 ±0.67; p=0.038; Fig. 3E) or ionomycin (AUC: 2.36±1.31 vs 1.20±0.56; p=0.022; Fig. 3F) following colchicine pre-treatment. Colchicine had no effect on spontaneous, PMA-or ionomycin-induced NET formation in neutrophils isolated from heathy participants (p=0.16, p=0.28, p=0.64, respectively; Supplementary Fig. 2).

### The effect of colchicine is independent of ROS signaling and ATP production

As the NET signalling cascade is largely dependent upon ROS generation, we assessed effects of colchicine on ROS in neutrophils derived from ACS patients. Colchicine had no effect on spontaneous (p=0.98; Fig. 3G), PMA-(p=0.38; Fig. 3H) or ionomycin-induced (p=0.96; Fig. 3I) ROS production; indicating that the effect of colchicine on NET formation was downstream of ROS generation.

To ensure the effect of colchicine was not a consequence of cellular toxicity, we measured ATP levels in neutrophils following colchicine treatment. Colchicine had no effect on cellular ATP production in either the presence or absence of stimulation, even up to the pre-defined time point of 2 h (Supplementary Fig. 3).

### Colchicine inhibits premature chromatin swelling in neutrophils isolated from patients with ACS

Downstream of ROS production, chromatin de-condensation and swelling acts as the point of no return for NET formation. As such, we directly visualized the effect of colchicine on this process. In the absence of stimulation there was no significant difference in chromatin area between heathy, ACS, untreated or colchicine-treated neutrophils (Fig. 4A and 4B; all p>0.6). After 1 h stimulation with PMA, there were clear differences between the groups (Fig. 4C and 4D). Neutrophils isolated from patients with ACS had 100% greater chromatin area compared to those isolated from healthy participants (103.7±34.9 vs 52.1±17.4 μm^2^; p<0.001). Colchicine pre-treatment resulted in a 35% reduction in the chromatin area of neutrophils isolated from patients with ACS (103.7±34.9 vs 66.8±39.0 μm^2^; p<0.001).

**Figure 4:**
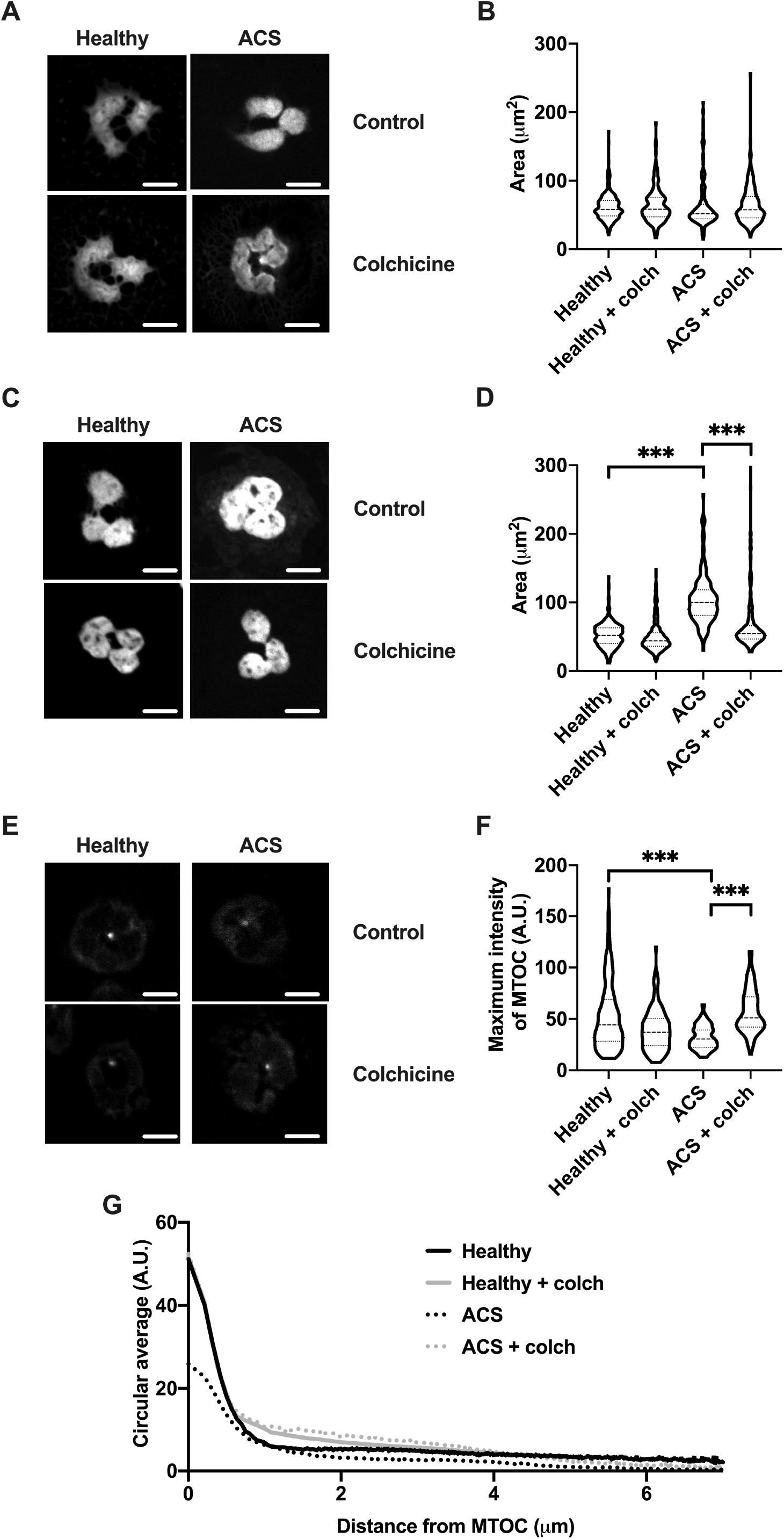
Colchicine inhibits premature chromatin swelling and restores α-tubulin organisation in neutrophils from patients with ACS. For quantification of chromatin swelling, neutrophils isolated from healthy participants and patients with ACS were treated with colchicine (colch; 25 nM) or vehicle control for 1.5 h. These cells were then fixed immediately (**A, B**) or stimulated with PMA (50 nM) for 1 h then fixed (**C, D**). All cells were stained with Hoechst prior to colchicine treatment and visualised by confocal microscopy. Chromatin area for individual cells was recorded in μm^2^ and compared between groups. For quantification of MTOC dynamics, neutrophils from healthy participants and patients with ACS were isolated, treated with colchicine (25 nM) or vehicle control for 1.5 h then fixed and stained with anti-α-tubulin and visualised by confocal microscopy. Maximum intensity of the MTOC (**E, F**) and staining intensity as a function of distance from the microtubule-organising centre (MTOC) was quantified for each condition (**G**). Scale bar = 5 μm. ***p < 0.005.

### Colchicine restores disrupted tubulin organization in neutrophils from ACS patients

A recent study demonstrated that rearrangement of the neutrophil cytoskeleton occurs during NETosis and may play a role in the early stages of NET release^24^. Under normal conditions, neutrophils have a well-defined MTOC that, upon activation, rearrange into structures resembling mitotic centrosomes. As colchicine markedly reduced chromatin swelling in ACS patient neutrophils, without altering ROS production, we hypothesized that colchicine-induced changes in cytoskeletal architecture may be responsible for halting NETosis. Neutrophils from healthy participants displayed a clear and well-defined MTOC (Fig. 4E). In contrast, α-tubulin organisation in neutrophils isolated from patients with ACS appeared to be disrupted, with a more diffuse staining pattern that was corrected by colchicine treatment. The MTOC maximal intensity was significantly reduced in neutrophils isolated from patients with ACS (32.16±32.73 A.U.) relative to both cells from healthy subjects (56.1±32.7 A.U.; p<0.005) and colchicine treated ACS neutrophils (57.94±21.50 A.U.; p<0.005). Quantification of α-tubulin staining intensity as a function of distance from the MTOC of each cell demonstrated close similarity between the signatures of healthy and colchicine treated ACS neutrophils, particularly within 1 μm of the MTOC (Fig. 4G).

## Discussion

This study demonstrates, for the first time, that colchicine acutely suppresses post-PCI local coronary release of key activated neutrophil-derived mediators in patients with ACS. Furthermore, we demonstrate that neutrophils isolated from ACS patients post-PCI are primed to undergo NETosis and that colchicine treatment acts directly on these cells to stabilize the cytoskeleton thereby, inhibiting NET release.

The clinical arm of this study identified several key findings. Firstly, untreated ACS patients had significantly higher baseline (pre-PCI) CS levels of all three activated neutrophil products (MPO, NE and NETs), and higher local release (post-PCI) of NETs and NE, compared to SAP patients. This indicates enhanced neutrophil activation and inflammatory mediator release from unstable plaques versus stable lesions at baseline and after PCI. Secondly, acute colchicine treatment markedly suppressed CS NETs and NE release post-PCI in ACS patients, compared to the untreated cohort. This suggests that colchicine acutely suppresses intra-plaque neutrophil activation after angioplasty-induced plaque disruption.

The *ex vivo* arm of this study was designed to investigate the direct effect of colchicine on neutrophils isolated from ACS patients post-PCI. These experiments yielded several novel findings. Neutrophils of ACS patients post-PCI have increased spontaneous and induced NET release, when compared to those of healthy control subjects. Low-dose colchicine treatment attenuates NET release from neutrophils isolated from ACS patients but not healthy controls. Quantification of α-tubulin staining indicates that this ‘selectivity’ of colchicine for ACS neutrophils may be related to microtubule disruption, which is evident in neutrophils isolated from ACS patients compared to healthy controls. At a cellular level, colchicine attenuated NET formation in a ROS-independent manner by preserving MTOC and α-tubulin integrity, thereby inhibiting chromatin swelling.

Neutrophil products are over-abundant in vulnerable coronary plaque and play a key role in peri-PCI MI in ACS patients. Purported mechanisms include PCI-induced disruption of the plaque-thrombus interface leading to release of pre-activated neutrophil mediators in vulnerable plaque and/or PCI-induced *de novo* neutrophil activation^19, 27^. In support of this, we previously reported a substantially higher release of neutrophil-derived microparticles and MPO into the coronary circulation after PCI of unstable lesions (versus stable lesions)^19^. Results of the present study corroborate these findings; PCI results in a marked release of NETs and neutrophils from ACS patients post-PCI are primed to undergo NETosis. This phenomenon is likely due to a combination of factors precipitated by PCI induced plaque disruption, such as increased pro-inflammatory cytokine levels, release of cholesterol crystals into the lumen and mechanical stress^10, 28, 29^. Priming of neutrophils lowers their threshold for NETosis, thereby predisposing individuals to a pro-inflammatory and pro-thrombotic cascade involving neutrophil-platelet aggregate formation and complement cascade activation, cytokine release, ROS generation causing oxidative damage, and local tissue degradation^3, 5-9^.

Colchicine is an inexpensive and widely available anti-inflammatory medication with an increasing evidence base demonstrating its efficacy in the secondary prevention of cardiovascular disease^15, 16, 30-32^. A small placebo controlled randomized trial showed lower cardiac enzyme release and reduced infarct size in colchicine treated STEMI patients versus controls^33^. It is well known that colchicine directly inhibits neutrophil chemotaxis^17^, although its effect on NET formation is less well defined. Two recent pre-clinical studies corroborate the inhibitory effect of colchicine on NET formation described in this study. First, *in vivo* colchicine administration limited cholesterol crystal-induced NETosis by inhibiting macropinocytosis – a microtubule-dependent process essential for mediating the pro-inflammatory effects of cholesterol crystals^34^. Second, *ex vivo* colchicine treatment of neutrophils isolated from individuals with Beçhet’s disease, was shown to markedly suppress spontaneous NETosis^35^. Adding to this, the present study demonstrates that colchicine inhibits NETosis in ACS patients by stabilizing the cytoskeleton, thereby attenuating chromatin swelling and subsequent NET release. Chromatin swelling is the major physical force driving NETosis and it has been shown to induce rupture of both the nuclear envelope and plasma membrane leading to extracellular DNA accumulation^24^. Furthermore, once initiated, chromatin swelling is irreversible and pharmacological inhibition of NETosis beyond this point is unlikely^24^. By limiting this process, colchicine provides a means of reducing NET-associated complications, particularly in inflammation-driven conditions.

This study examines neutrophil activation products in the CS during PCI and circulating neutrophils post-PCI. It is important to consider that NETs, MPO and NE measured in the CS during PCI were largely a result of intraplaque neutrophil activation, induced by a combination of localized inflammation and direct mechanical stress^36^. Nevertheless, *in vitro* data from this study further demonstrates systemic neutrophil ‘priming’ post-PCI. Although these circulating neutrophils are not terminally activated to undergo NETosis, they demonstrate enhanced reactivity to stimuli – a ‘primed’ state^37,38^.We show that colchicine not only attenuates the localized release of neutrophil activation products in the coronary circulation but also that colchicine promotes transition of circulating ‘primed’ neutrophils back to a basal state.

By utilising *in vivo* and *ex vivo* experimental techniques, we demonstrate a robust effect of colchicine on neutrophil activation in the peri-procedural period. Nonetheless, there are several important limitations of this study. First, the relatively small sample size and non-randomized nature of the allocation process. Despite this, the treatment and control groups were statistically well matched at baseline. Secondly, intra-vascular imaging was not used to confirm that plaque morphology corresponded with clinical presentation. Thirdly, there were more patients with three-vessel disease and six higher risk PCIs performed in the treatment group. This could represent an imbalance in the potential neutrophil activation kinetics between the two groups, although it would likely skew the results to favor the efficacy of colchicine even further. Finally, despite showing a clear reduction in neutrophil-mediated inflammatory mediator release post-PCI with colchicine treatment, this study was not powered to test harder clinical endpoints, such as microvascular obstruction or peri-procedural MI.

In ACS patients who underwent PCI, colchicine acutely and markedly suppressed local neutrophil activation by directly inhibiting NET release from primed neutrophils. These results provide further evidence of the beneficial role of colchicine in patients with ACS, particularly in the peri-procedural period. Larger randomised trials powered for hard clinical outcomes are required to evaluate whether colchicine is effective at reducing peri-procedural MI and MACE in ACS patients.

## Data Availability

Data is available from the corresponding authors by request

## Abbreviations

ACS: Acute coronary syndrome
AUC: Area under the curve
CS: Coronary sinus
MACE: Major adverse cardiovascular event
MI: Myocardial infarction
MPO: Myeloperoxidase
MTOC: Microtubule organisation centre
NE: Neutrophil elastase
NET: Neutrophil extracellular trap
PCI: Percutaneous coronary intervention
PMA: Phorbol 12-myristate 13-acetate
ROS: Reactive oxygen species
SAP: Stable angina pectoris

## Acknowledgements

The authors thank the cardiac catheterization laboratory staff at the Royal Prince Alfred Hospital and the Biomedical Imaging Facility staff at UNSW Sydney for their assistance in performing these studies.

## Highlights

- Patients with ACS have enhanced neutrophil activity relative to patients with stable angina pre-and post-PCI.
- Colchicine treatment significantly reduced the production of NETs in ACS patients *in vivo*.
- Neutrophils isolated from ACS patients are primed to undergo NETosis, and this is inhibited by colchicine.
- Microtubule organisation is impaired in neutrophils isolated from patients with ACS patients, but is restored by treatment with colchicine.

